# Spectrum of Genetic Disorders and Gene Variants in the United Arab Emirates National Population: Insights from the CTGA Database

**DOI:** 10.1101/2023.03.03.23286732

**Authors:** Sami Bizzari, Pratibha Nair, Sayeeda Hana, Asha Deepthi, Mahmoud Taleb Al-Ali, Lihadh Al-Gazali, Stephany El-Hayek

**Affiliations:** Centre for Arab Genomic Studies, Dubai, United Arab Emirates; Department of Pediatrics, College of Medicine and Health Sciences, United Arab Emirates University, Al Ain, United Arab Emirates

**Keywords:** variant, Emirati, database, variant annotation, rare disease

## Abstract

Like many other Arab countries, the United Arab Emirates (UAE) has a relatively high prevalence of genetic disorders. Here we present the first review and analysis of all genetic disorders and gene variants reported in Emirati nationals and hosted on the Catalogue for Transmission Genetics in Arabs (CTGA), an open-access database hosting bibliographic data on human gene variants associated with inherited or heritable phenotypes in Arabs. To date, CTGA hosts 665 distinct genetic conditions that have been described in Emiratis, 621 of which follow a clear Mendelian inheritance. Strikingly, over half of these are extremely rare according to global prevalence rates, predominantly with an autosomal recessive mode of inheritance. This is likely due to the relatively high consanguinity rates within the Emirati population. The 665 conditions include disorders that are unique to the Emirati population, as well as clearly monogenic disorders that have not yet been mapped to a causal genetic locus. We also describe 1,365 gene variants reported in Emiratis, most of which are substitutions and over half are classified as likely pathogenic or pathogenic. Of these, 235 had not been reported on the international databases dbSNP and Clinvar, as of December 2022. Further analysis of this Emirati variant dataset allows a comparison of clinical significance as reported by Clinvar and CTGA, where the latter is derived from the study cited. A total of 307 pathogenic/likely pathogenic variants from CTGA’s Emirati dataset, were classified as benign, variants of uncertain significance, or were missing a clinical significance or had not been reported by Clinvar. In conclusion, we present here the spectrum of genetic disorders and gene variants reported in Emiratis. This review emphasizes the importance of ethnic databases such as CTGA in addressing the underrepresentation of Arab variant data in international databases and documenting population - specific discrepancies in variant interpretation, reiterating the value of such repositories for clinicians and researchers, especially when dealing with rare disorders.

## Introduction

The UAE is made up of seven Emirates that collectively cover an area of 83,600 km ^2^. The demographic of the UAE is unique in that close to 90% of the population is expatriate (CIA, 2023).

UAE nationals, or Emiratis, that make up about 11% of the population, are an ethnically diverse admixture of Middle Eastern, African and South Asian ancestries, resulting from various human migrations that have crossed along the Arabian Peninsula (Alshamali et al., 2009 ; Aljasmi et a l., 2020; Tay et al., 2020). In this review we focus on the landscape of genetic disorders and their associated gene variants, exclusively in Emiratis.

As is the case in many other Arab countries, there is a relatively high prevalence of genetic disorders in the UAE. A few reports, though not recent, paint a very clear picture of this. The 2006 March of Dimes report ranks the UAE 6^th^ in the world in terms of prevalence of genetic and partially genetic birth defects (Arnold Christianson, 2006). Hospital based studies reported that 0.31% of live births had neonatally lethal anomalies while 6.9% had congenital anomalies (Dawodu et al., 2005; Aryasinghe et al., 2012). These high rates are largely due to the tribal nature of Emirati culture with an elevated consanguinity rate (50%) and the presence of isolated populations with increased levels of inbreeding. Other contributing factors include large family size as well as high maternal and paternal ages at conception (Al-Gazali and Ali, 2010). Consequently, the Emirati genome, like other Arab genomes, carries significantly enriched regions of homozygosity, which in turn contributes to the higher burden of recessive genetic disorders (Alkuraya, 2010).

In the last decade, the UAE has greatly expanded its healthcare infrastructure, including its genetic services. The number of genetic centers has increased making diagnostic testing and the essential counseling services more widely available. This has also made testing more affordable and faster (Abou Tayoun et al., 2021). Several research centers have also been established which partly accounts for the tremendous increase in genetic and genomic Emirati data being generated. The Government’s prioritization of investment in the fields of genetics and genomics has also resulted in the launch of the Emirati Genome Project which is currently underway (Al-Ali et al., 2018).

Ethnic and/or national genetic databases offer an essential tool for clinicians, and are also a valuable asset that can direct screening programs (premarital, prenatal, and newborn) as well as national health priorities. The latter is accomplished by such databases that provide a clear snapshot of the current landscape entailing genetic disorders in a region, while simultaneously highlighting relatively common genetic disorders and disease-causing variants specific to the unique genetic architecture of the concerned populations (Patrinos, 2006). The Catalogue for Transmission Genetics in Arabs (CTGA) is an open-access database hosting bibliographic data on human gene variants associated with inherited or heritable phenotypes in Arabs (www.cags.org.ae/ctga). This data is particularly relevant, considering the tremendous gap and under-representation of Arabs in international databases. GnomAD’s 158 middle eastern genomes make up only about 0.2% of its latest release v3.1 (Abou Tayoun and Rehm, 2020). The figure is more extreme for GWAS studies, where up to 2021, Arabs constituted 0.01% of total participants (Mills and Rahal, 2020). There is thus a pressing need for better access and more comprehensive coverage of genetic data on Arabs. Although there have been attempts to fill this gap by other databases (Scott et al., 2016; Koshy et al., 2017; Karczewski et al., 2020; Vatsyayan et al., 2021), to the best of our knowledge, CTGA is the most comprehensive compendium of bibliographic genetic data on Arab populations.

The latest review of genetic disorders in the UAE (which included Emiratis and non-Emiratis), albeit not recent, lists 270 types of disorders, the majority of which are rare autosomal recessive single gene disorders (Al-Gazali and Ali, 2010). Here we present the first review and analysis of all genetic disorders and gene variants reported in Emirati nationals and hosted on CTGA.

## Materials and Methods

### Literature Review

A comprehensive literature search was conducted for biomedical literature originating from the UAE or referring to Emirati subjects from 2010 to 2021 on PubMed using the search clause “*UAE OR Emirat\** “. Additionally, Harzing’s Publish or Perish V8 software was used to extract citations from Google Scholar with a limitation of 1,000 articles per year using the following refined search clause: “*UAE OR Emirati OR Emirate mutation OR variant OR gene OR genetic patient OR individual OR subject OR cohort OR proband OR case”*. Terms that would populate irrelevant results, such as “Ubiquitin Activating Enzyme” for “UAE”, were filtered out in the search string.

Obtained articles were manually screened with the following inclusion criteria: 1. Articles describing a genetic disease or phenotype in an Emirati individual(s) and 2. Articles describing genetic variants in an Emirati individual(s) or in the Emirati population as a whole. The following exclusion criteria were maintained: 1. Articles with potentially relevant data where the ethnicity of the subject(s) could not be confirmed in the article or by the authors once contacted and 2. Articles with redundant data including reviews and duplicate information.

### Data Curation

Relevant information from these articles was curated and extracted onto CTGA (cags.org.ae/ctga), a SQL relational database where data is arranged primarily by anonymous subjects, concisely detailing their clinical features, diagnosed conditions, and associated genetic variants and bibliographic sources. The various categories of data extracted from each article included the number of subjects or patients, sex, presence of parental consanguinity and/or family history, clinical features, diagnosed condition, any molecular (genetic variant) diagnosis, bibliographic citation, as well as molecular details of healthy individuals (such as relatives of probands or control groups), if available. Anonymized subject and family IDs were given for each distinct subject or subject group entered into the database.

Extracted data was converted to universally recognized descriptions using online API sources. Terms for subject clinical features were derived from the Human Phenotype Ontology (HPO) (hpo.jax.org). Associated condition and gene records were linked to their corresponding OMIM records (omim.org) and classified with WHO-ICD wherever available. Variant HGVS terms were retrieved using Variant Validator (variantvalidator.org) and/or Ensembl (ensembl.org) and linked to relevant subjects along with their dbSNP and ClinVar IDs (ncbi.nlm.nih.gov) whenever available. Variant types were based on classification by VarNomen (varnomen.hgvs.org). Variant zygosity, its clinical significance according to ClinVar, and allele counts were recorded for each subject, as well as allele frequencies for large groups of individuals in association studies. The clinical significance derived from the literature was entered as CTGA clinical significance, for each variant. Data on variants not compatible with HGVS terms, as well as articles reporting only clinical data, were recorded as text-based descriptions. Bibliographic sources were linked to subjects with their PubMed ID (or link to journal page if not indexed) and reference title cataloged in the NLM format (ncbi.nlm.nih.gov).

### Comparison with other Databases

The Emirati variant dataset hosted on CTGA was checked for availability on two international genomic databases, namely dbSNP and ClinVar. In order for us to compare how clinical significance of Emirati variants is reported on CTGA vs Clinvar, we considered only variants without conflicting significances and obtained a list of variants where CTGA clinical significance (CS) is described as “likely pathogenic” and/or “pathogenic”, while Clinvar describes the same variants as “benign”, “likely benign”, “uncertain significance” or does not report their CS. Data was last updated on the 29^th^ of December, 2022.

## Results

Out of 15,599 surveyed articles, 334 relevant publications reporting genetic data on Emirati subjects and published between 2010 and 2021 were curated, which when added to existing papers from previous surveys, brought the total of UAE-related papers on CTGA to 487. Adding the relevant data from these papers increased the number of Emirati subjects on the CTGA database, to a total of 11,875 subjects, diagnosed with 665 different genetic disorders/phenotypes. The number of Emirati individuals for whom data is available on CTGA is actually higher as this figure does not account for subjects lacking molecular data (which were curated from clinical studies), unaffected relatives, or control subjects. In addition, while dealing with studies where the actual number of patients was not specified, only the minimum confirmable number of patients was added to the total count.

As mentioned above, access to CTGA is free, and users are encouraged to carry out their own data analyses. We provide here some of the various analyses of the Emirati data in the CTGA Database.

### Noteworthy phenotypes

A comprehensive list of the 665 genetic phenotypes reported in Emirati subjects is available online (Supplementary Table 1). Table 1 lists a subset of these diseases that warrant a special mention, in that they have been reported so far exclusively in Emirati patients and/or were first mapped in Emirati families. Ten of these disorders are exclusive to Emirati patients, eight of which are rare autosomal recessive disorders reported in consanguineous families. Also included in Table 1 are the five genetic disorders that to date have not been assigned to a specific gene locus as well as eight disorders with novel gene-phenotype correlations that have not yet been reported on OMIM. The latter category includes disorders such as Nonsyndromic Macular Pseudocoloboma, *ITFG2* Associated Neurodevelopmental Disorder, and *BMPR1A*-Related Syndrome. In addition to these novel associations listed in Table 1, the database also documents neurodevelopmental disorders with unconfirmed causal gene associations, such as those reported for *MAP3K9* and *ASB11* (Saleh et al., 2021) as well as large chromosomal duplication and deletion syndromes (Alabdullatif et al., 2017).

**Table 1.**
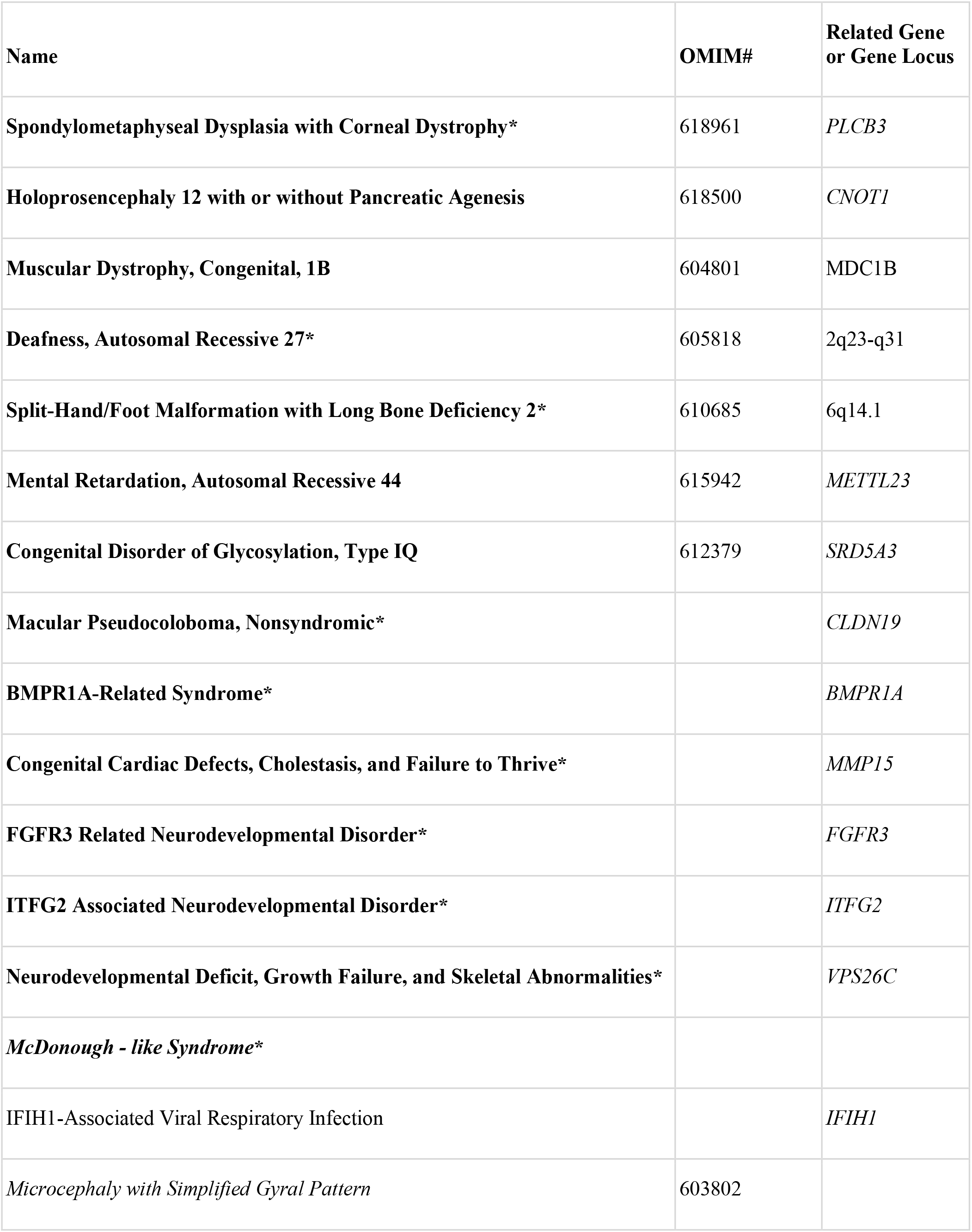
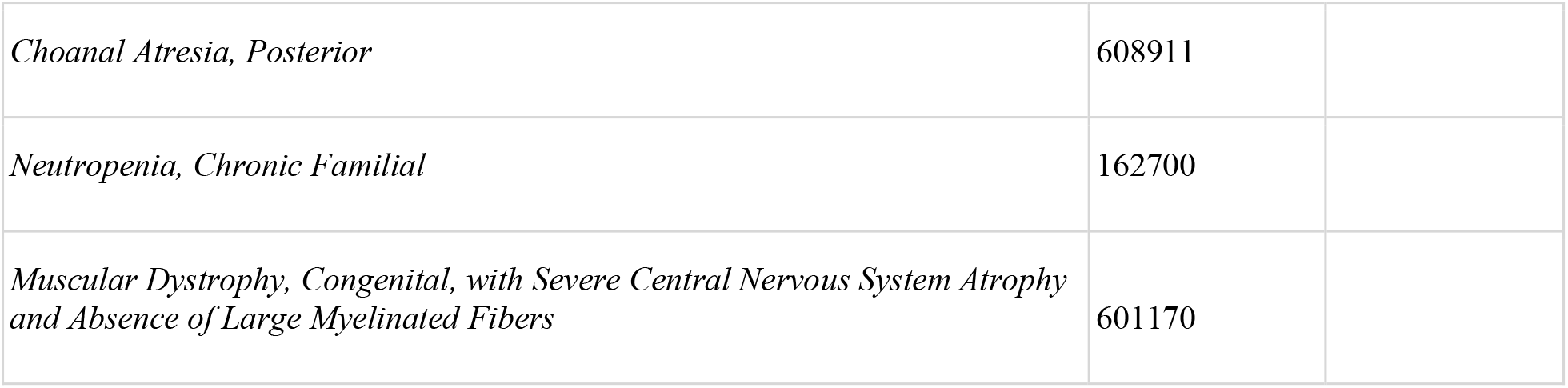
Selected disorders from the list of genetic phenotypes reported in Emirati subjects. Disorders with names in bold were first mapped or described in Emirati families. * denotes disorders described only in Emiratis to date. Names in italics indicate unmapped disorders where familial cases have been reported on CTGA.

A major part of these genetic disorders reported in the UAE are rare in their distribution, based on worldwide prevalence rates provided by Orphanet (https://www.orpha.net/ Fig. 1). Around 55% of these disorders are classified as having a prevalence of <1 in 100,000 individuals, while an additional 13% fall in the prevalence range of 1-9/100,000 individuals. In addition, 20% of these disorders could not be classified according to global prevalence rates. These include disorders such as beta - thalassemia that are highly prevalent in the Arab World, but are rare globally, as well as specific sub - types of relatively common genetic disorders, such as Developmental and Epileptic Encephalopathy or Autosomal Recessive Deafness.

**Figure 1:**
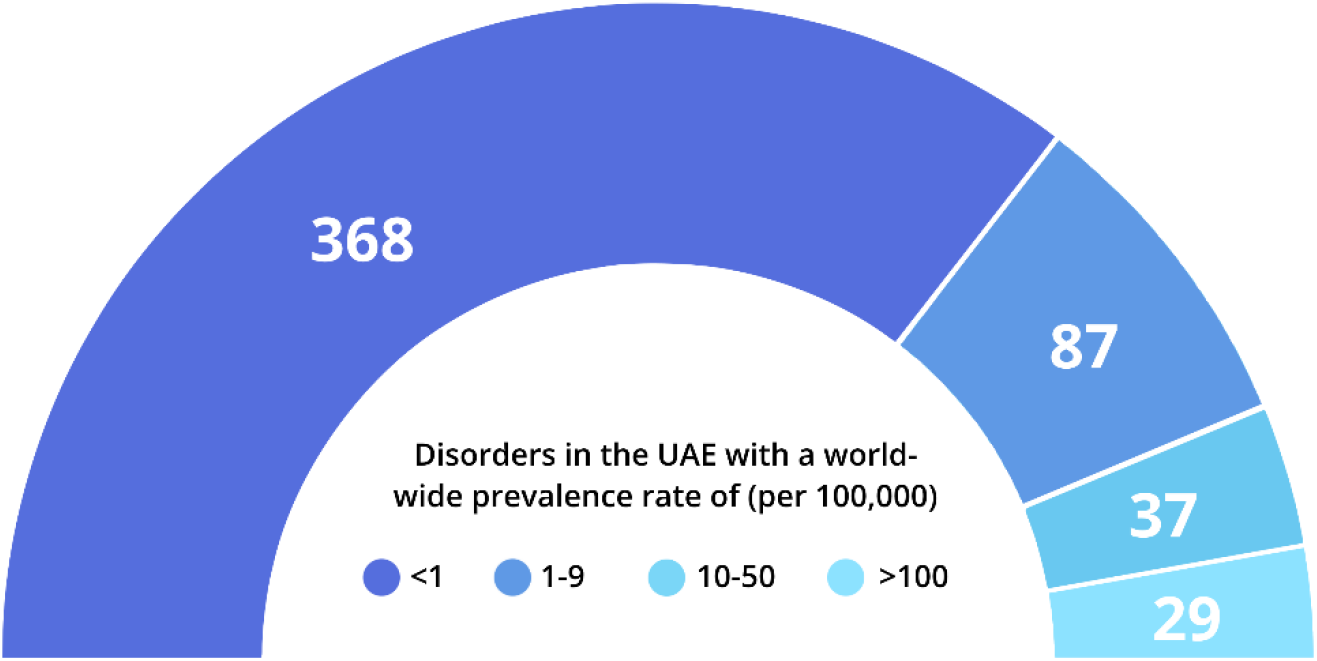
Distribution of genetic disorders in the UAE according to available world-wide prevalence rates. Prevalence rates were obtained from Orphanet (orpha.net) where available. 135 disorders lacked sufficient prevalence data; 9 phenotypes described in UAE individuals (e.g., blood groups; slow acetylation) were not included in this representation.

### Non-multifactorial disorders

The majority of phenotypes/conditions reported in Emiratis on CTGA (93.4%) follow a purely genetic inheritance pattern (referred to as non-multifactorial from here onwards) whereas multifactorial disorders that carry a genetic component, such as Type 2 Diabetes Mellitus, Hypertension, and Metabolic Syndrome make up a smaller fraction. A detailed analysis of the non - multifactorial conditions is presented in the following sections.

A closer look at the number of families reported for each of the non-multifactorial disorders, whereby all related individuals count as a single family, expectedly highlights disorders such as Cystic

Fibrosis (OMIM# 219700), Beta-Thalassemia (OMIM# 613985), and Sickle Cell Anemia (OMIM# 603903), which have been reported in at least 30 unrelated Emirati families and shown to be highly prevalent in the UAE (Elsaban et al., 2021). Moreover, we note the striking occurrence of several globally rare diseases, such as Stuve-Wiedemann Syndrome (OMIM# 601559), Stargardt Disease 1 (OMIM# 248200), Biotinidase Deficiency (OMIM# 253260), and Phenylketonuria (OMIM# 261600) in more than ten distinct Emirati families, as well as some rare disease subtypes like Joubert Syndrome 3 (OMIM# 608629), Autosomal Recessive Deafness Type 1A (OMIM# 220290), and Lysosomal Mannosidosis Type Alpha B (OMIM# 248500), in at least four unrelated Emirati families, each. In fact, 18 of the 37 non-multifactorial disorders which have been reported on CTGA in at least four Emirati families have a global prevalence of <1 in 100,000 individuals (Table 2).

**Table 2:**
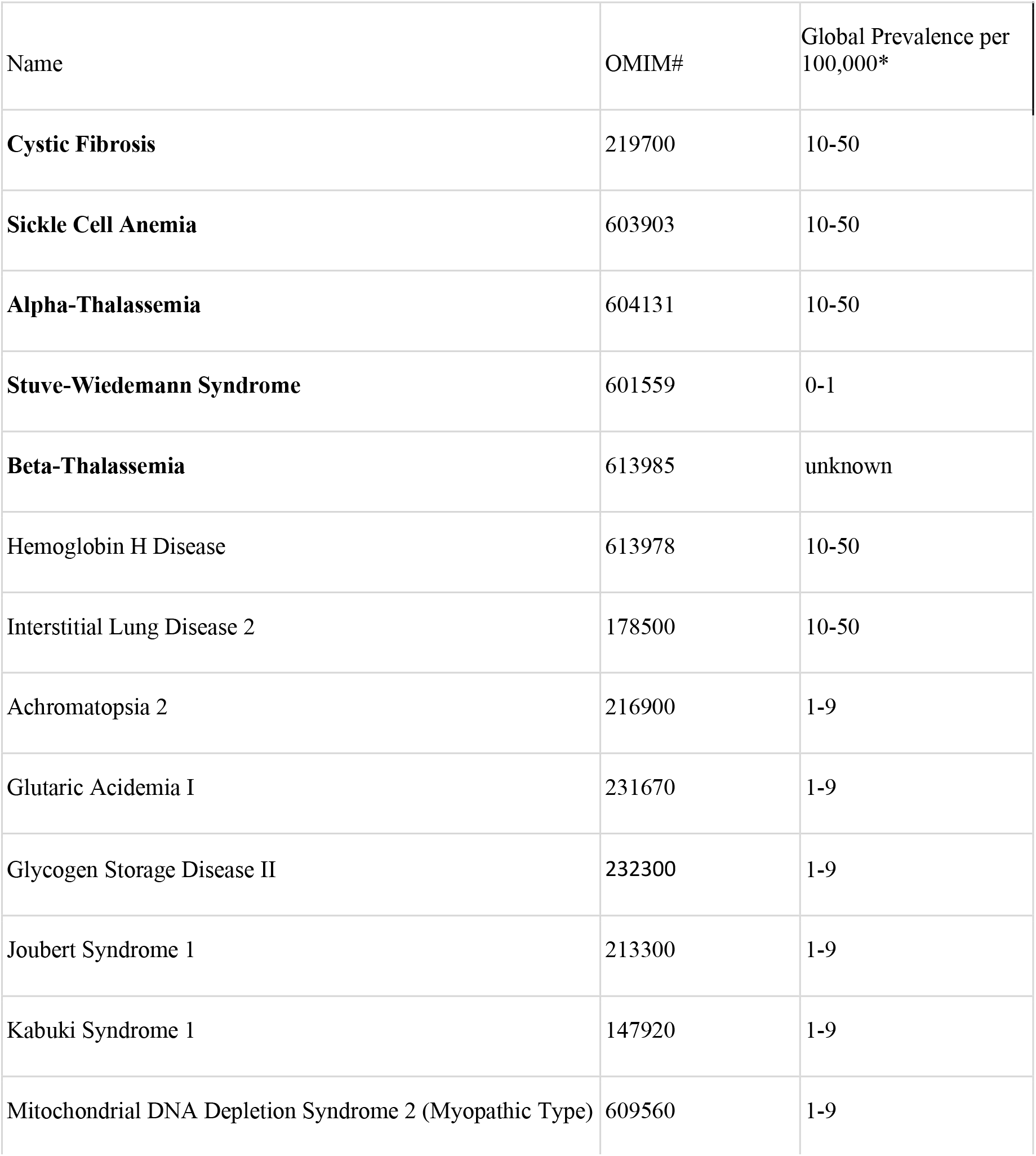
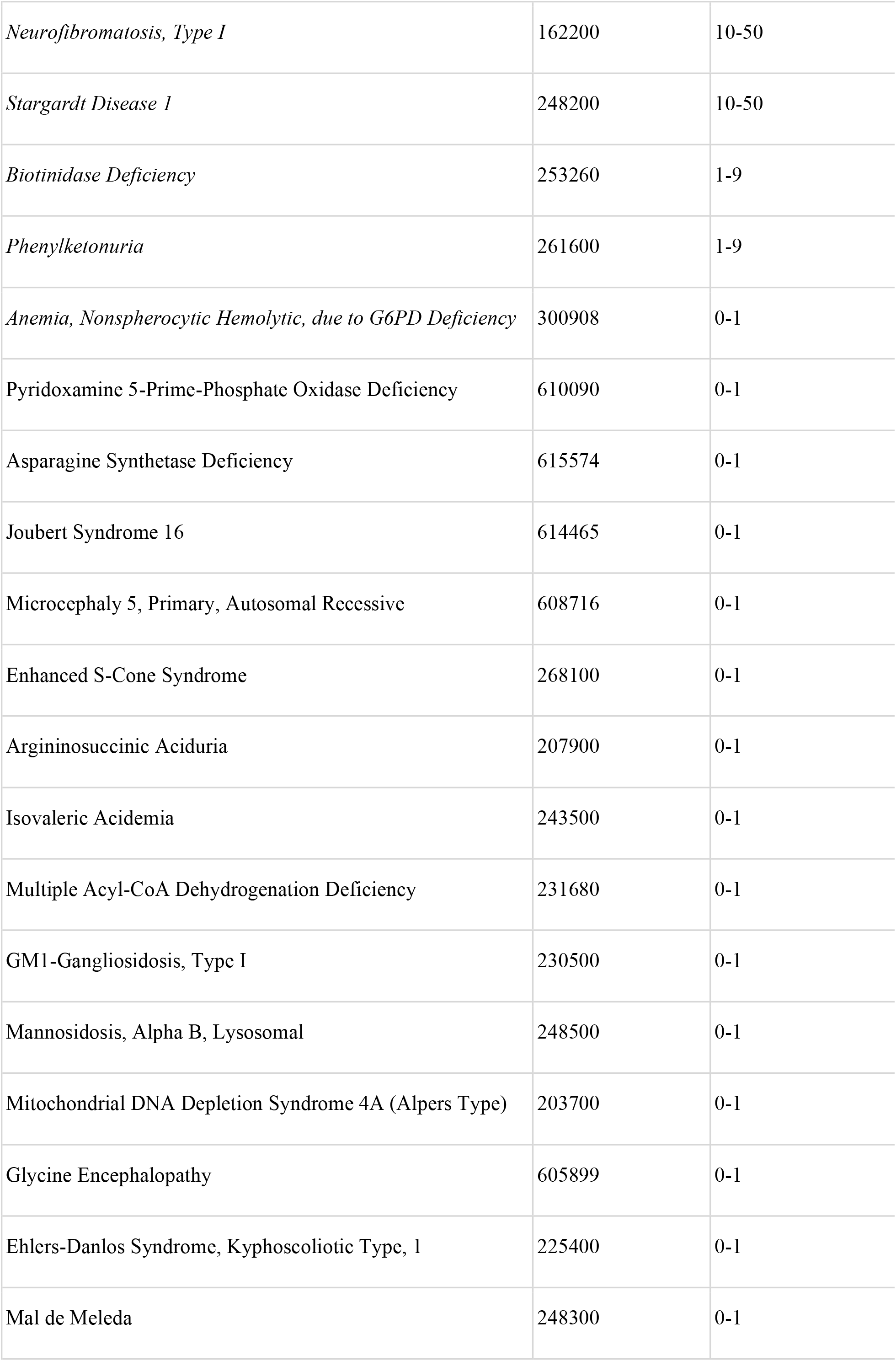
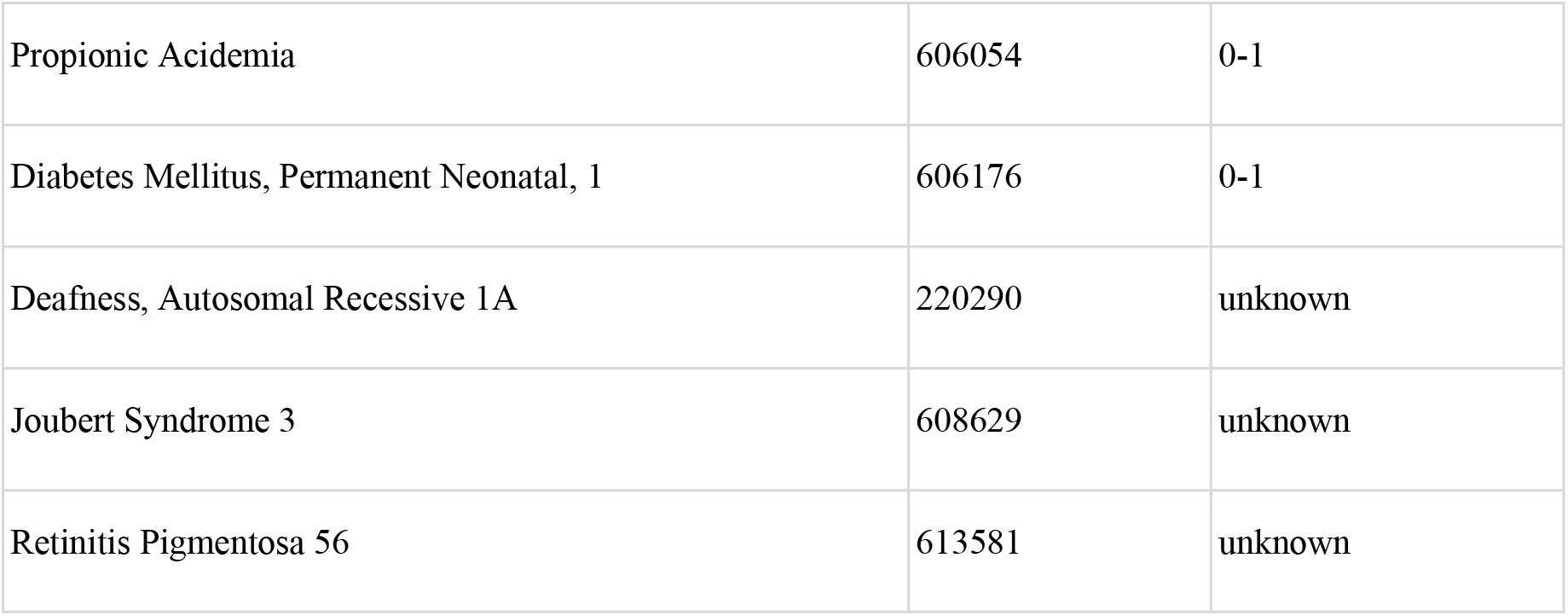
Disorders reported in at least four Emirati families. Disorders in bold have been described in at least 30 families. Disorders in italics have been described in at least 10 families. *Global prevalence is based on Orpha.net.

| Name | OMIM# | Global Prevalence per 100,000* |
| --- | --- | --- |
| **Cystic Fibrosis** | 219700 | 10-50 |
| **Sickle Cell Anemia** | 603903 | 10-50 |
| **Alpha-Thalassemia** | 604131 | 10-50 |
| **Stuve-Wiedemann Syndrome** | 601559 | 0-1 |
| **Beta-Thalassemia** | 613985 | unknown |
| Hemoglobin H Disease | 613978 | 10-50 |
| Interstitial Lung Disease 2 | 178500 | 10-50 |
| Achromatopsia 2 | 216900 | 1-9 |
| Glutaric Acidemia I | 231670 | 1-9 |
| Glycogen Storage Disease II | 232300 | 1-9 |
| Joubert Syndrome 1 | 213300 | 1-9 |
| Kabuki Syndrome 1 | 147920 | 1-9 |
| Mitochondrial DNA Depletion Syndrome 2 (Myopathic Type) | 609560 | 1-9 |
| *Neurofibromatosis, Type I* | 162200 | 10-50 |
| *Stargardt Disease 1* | 248200 | 10-50 |
| *Biotinidase Deficiency* | 253260 | 1-9 |
| *Phenylketonuria* | 261600 | 1-9 |
| *Anemia, Nonspherocytic Hemolytic, due to G6PD Deficiency* | 300908 | 0-1 |
| Pyridoxamine 5-Prime-Phosphate Oxidase Deficiency | 610090 | 0-1 |
| Asparagine Synthetase Deficiency | 615574 | 0-1 |
| Joubert Syndrome 16 | 614465 | 0-1 |
| Microcephaly 5, Primary, Autosomal Recessive | 608716 | 0-1 |
| Enhanced S-Cone Syndrome | 268100 | 0-1 |
| Argininosuccinic Aciduria | 207900 | 0-1 |
| Isovaleric Acidemia | 243500 | 0-1 |
| Multiple Acyl-CoA Dehydrogenation Deficiency | 231680 | 0-1 |
| GM1-Gangliosidosis, Type I | 230500 | 0-1 |
| Mannosidosis, Alpha B, Lysosomal | 248500 | 0-1 |
| Mitochondrial DNA Depletion Syndrome 4A (Alpers Type) | 203700 | 0-1 |
| Glycine Encephalopathy | 605899 | 0-1 |
| Ehlers-Danlos Syndrome, Kyphoscoliotic Type, 1 | 225400 | 0-1 |
| Mal de Meleda | 248300 | 0-1 |
| Propionic Acidemia | 606054 | 0-1 |
| Diabetes Mellitus, Permanent Neonatal, 1 | 606176 | 0-1 |
| Deafness, Autosomal Recessive 1A | 220290 | unknown |
| Joubert Syndrome 3 | 608629 | unknown |
| Retinitis Pigmentosa 56 | 613581 | unknown |

We also analysed the distribution of the modes of inheritance of genetic conditions reported in Emiratis (Fig.2A). Nearly two-thirds of all non-multifactorial conditions reported in the UAE were listed as having an exclusively autosomal recessive (AR) mode of inheritance on OMIM. This was followed by autosomal dominant and X-linked disorders. A significant number of disorders are classified as having multiple modes of inheritance (Supplementary Table 1). For example, CTGA describes the AR transmission of Dopa-Responsive Dystonia (OMIM# 128230), a predominantly dominant disorder, in an Emirati subject exhibiting a recessive mode of transmission.

**Figure 2:**
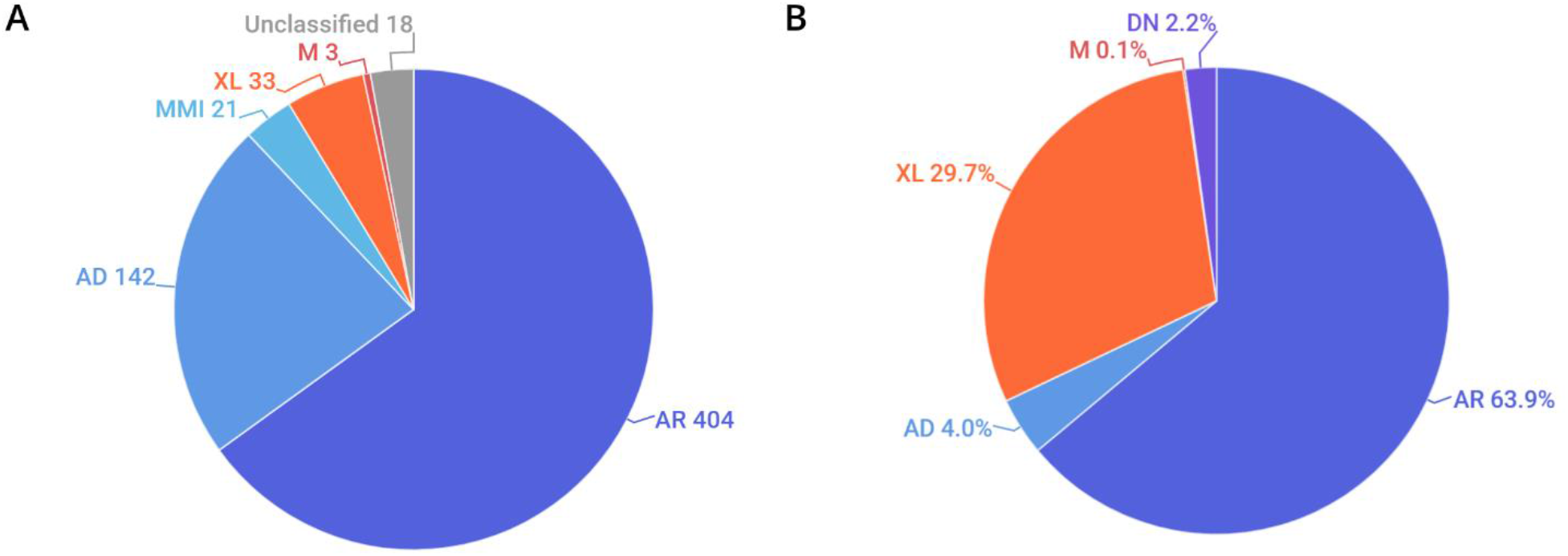
Distribution of the mode of inheritance by (**A)**. number of disorders and **(B**). percentage of Emirati subjects. AR, autosomal recessive; AD, autosomal dominant; MMI, multiple modes of inheritance; XL, X-linked; M, mitochondrial; DN, de novo. Unclassified conditions include multifactorial disorders, chromosomal disorders, susceptibility to certain disorders, as well as non - disease phenotypes.

Examining the distribution of modes of inheritance according to subjects showed a similar trend, where the AR mode of transmission was associated with the largest proportion (63.9%) of Emirati patients reported on CTGA. Notably, among patients with AR transmission for whom parental consanguinity status was provided, over 95% exhibited some degree of consanguinity. X-linked disorders, which account for about 29.7% of Emiratis on CTGA, were the next largest category of disorders recorded. This large ratio can be attributed to the high number of Emirati subjects reported with G6PD deficiency, which exhibits relatively high prevalence rates in the UAE (Elsaban et al., 2021) (Fig.2B). Additionally described on CTGA are Emirati subjects with *de novo* variants; these are mostly associated with dominant (43 of 46) disorders, such as Kabuki Syndrome type I (OMIM# 147920), for which we report five separate *de novo* variants. Other *de novo* variants described involved sporadic somatic mosaicism, including Emirati patients with Schimmelpenning-Feuerstein-Mims Syndrome (OMIM# 163200), as well as a sporadic mutation in a male patient with X-linked SCID (OMIM# 300400).

One of the most significant benefits of the CTGA database to clinicians is that it contains detailed phenotypic data for patients and patient groups, curated in the form of Human Phenotype Ontology (HPO) terms. Supplementary Table 2 lists the 20 most common HPO terms reported in Emirati patients. The most common term was found to be ‘anemia’, which can be attributed to the large number of patients with hemoglobinopathies, a prevalent class of disorders in the UAE. Neurodevelopmental phenotypes, such as ‘global developmental delay’, ‘seizure’, ‘hypotonia’, and intellectual disability were also found to be commonly represented; a fact further supported by the high numbers of ‘disorders of the nervous system’ seen when we classify the 665 disorders according to WHO-ICD10 chapters (Supplementary Table 1).

### Variants

CTGA currently describes a total of 1,365 variants in 844 genes reported in Emirati subjects; over 63% of these variants are associated with non-multifactorial conditions. A complete list of the 1,365 variants can be downloaded from https://cags.org.ae/en/search-database. The large majority of these variants are substitutions (74%), followed by deletions (10%), and reference alleles (6%) (Fig.3). A comparison with datasets from dbSNP and Clinvar, reveals that as of December 2022, 235 (17.2%) of all UAE variants on CTGA were exclusively reported on our database and thus absent from both these databases. An additional 446 variants were available on dbSNP, but not on Clinvar. The latter includes 55 reference alleles reported with significant associations with common disorders such as Type 2 Diabetes Mellitus and Obesity.

**Figure 3:**
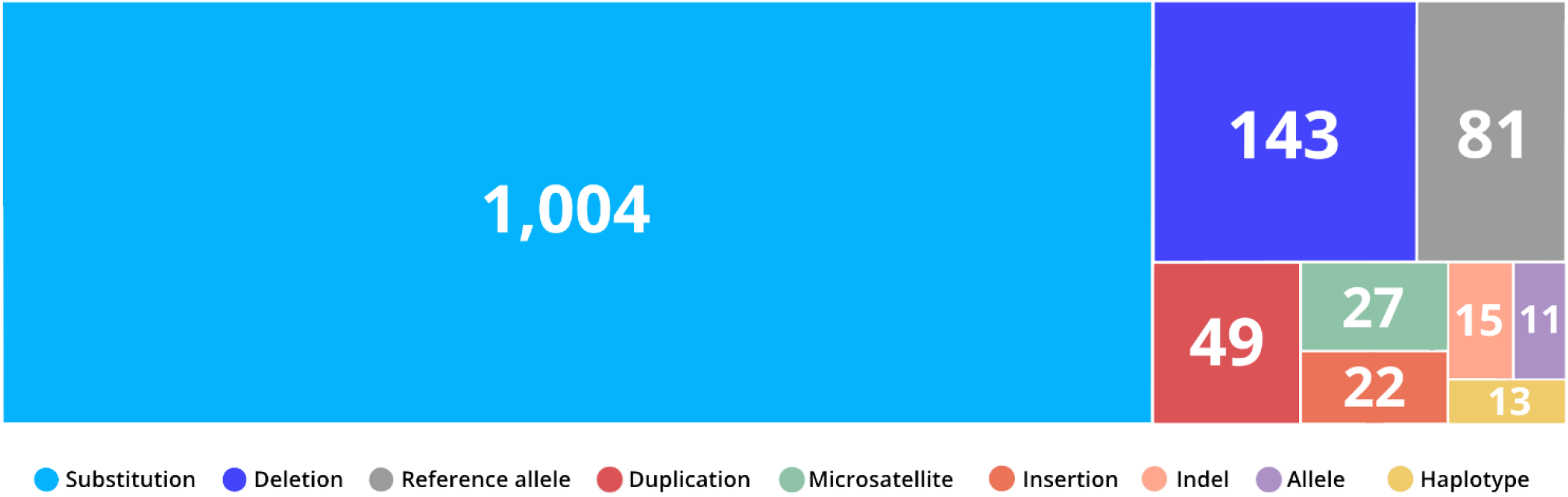
Distribution of variant types reported in the UAE. Variants were classified according to VarNomen. Reference alleles were derived from dbSNP.

The clinical significance of a gene variant is one of the main components of the diagnostic pipeline for genetic disorders (Sundercombe et al., 2021). On CTGA, we report CS according to Clinvar, where available, as well as CTGA CS. The latter is derived from the findings and variantanal yses described by the paper(s) where subject(s) harboring the variant were described. Around 54% (739) of Emirati variants on CTGA were classified as pathogenic or likely pathogenic by at least one of the reporting papers. A total of 99 variants were identified unanimously as variants of uncertain significance in the studies where they were reported. These variants require reannotation as more evidence comes to light.

Interestingly, many variants show a marked difference between the CS reported on ClinVar and CTGA. When considering variants we classify as exclusively likely pathogenic or pathogenic based on the findings of the reporting studies, we find that 55 were classified as benign variants, variants of uncertain significance (VUS), or lacked a clinical significance on Clinvar. An additional 252 had not been reported by Clinvar. Of these 307 variants, 41 were identified on the gnomAD v3.1.2 database (gnomad.broadinstitute.org) exhibiting an allele frequency below 1%, and 17 were identified on the GME variome database (igm.ucsd.edu/gme/) with an allele frequency below 1% (Last updated in December 2022).

## Discussion

CTGA is a continuously growing, manually curated database. As of March 2023, CTGA holds data on 2,297 genetic diseases and phenotypes and 4,156 variants reported in Arab individuals. This allows our database to not only provide a depiction of the genetic conditions and their causal gene variants in each Arab country, but also define the landscape of such conditions across the Arab ethnic group. In the current study, we provide a comprehensive qualitative and quantitative assessment of the spectrum of genetic disorders and their associated gene variants in the Emirati population.

In terms of healthcare, UAE currently faces a dual challenge; firstly from the preponderance of non - communicable diseases, such as diabetes, cancers, cardiovascular conditions, and obesity (Elmusharaf et al., 2022; Fadhil et al., 2022), all of which have a pronounced genetic component, and secondly from the presence of common and rare monogenic disorders. This is reflected in two metrics of the Emirati data within CTGA; the number of reported disorders and the number of reported Emirati subjects. Interestingly enough, even though most unique disorders on the database are purely genetic and rare (Fig.1), multifactorial disorders account for a large number of our subjects. For example, diabetes, hypertension, and asthma cover over 45% of Emirati patients on CTGA. The actual number of disorders described in Emiratis is likely higher than the 665 phenotypes that we report, as this figure does not take unpublished data into account. A similar review was made more than a decade ago (Al-Gazali and Ali, 2010), and although smaller in scale, it had documented 270 distinct monogenic diseases in the UAE. This increase is mainly due to the enhanced ability to diagnose genetic disorders and a consequent surge in the reporting of such disorders within the past decade. With the advent and ease of access to technologies such as Whole Exome Sequencing (WES) and gene panel sequencing, diagnostic rates of genetic disorders have increased significantly (Vaisitti et al., 2021 ; Sanchez-Luquez et al., 2022; Moundir et al., 2023). In fact, within our set of genetic disorders reported in the UAE, a significant portion comes from the utilization of such sequencing techniques on fairly large patient populations (Al-Shamsi et al., 2016; Alsamri et al., 2020 ; Mahfouz et al., 2020; Saleh et al., 2021). Additionally, the UAE has recently prioritized research on clinical genetics and the inclusion of genetics in the country’s health priorities. A cursory analysis of the number of publications on PubMed using the search terms “UAE” and “genetic” clearly shows this increase in research output. Compared to 277 publications on the database up until 2010, there has been a more than 400% increase in the number of publications since 2010. In fact, the research output for clinical genetics in the UAE was double the global rate in 2018 (MOHAP, 2019).

Consanguineous unions are known to be at a higher risk of producing offspring with birth defects and inherited disorders (Majeed-Saidan et al., 2015; Ben-Omran et al., 2020). In addition, consanguinity and consequent homozygosity have also been shown to play a role in the etiology of complex diseases (Bittles and Black, 2010; Fareed and Afzal, 2017). In previous studies on other Arab populations, we have shown increasing proportions of recessive disorders with high rates of consanguinity (Bizzari et al., 2018; Bizzari et al., 2021). The high rate of consanguinity and increased inbreeding coefficient within the Emirati population (al-Gazali et al., 1997) is likely to be one of the major factors underlying the large proportion of recessive disorders that we report. This is substantiated by the fact that the majority of Emiratis diagnosed with recessive disorders in the database are from consanguineous unions. The breakdown of disease inheritance patterns in individual patients further emphasizes this association, with a significantly large proportion of patients affected by recessive disorders; these particularly include hemoglobinopathies such as Alpha-Thalassemia, Beta-Thalassemia, and sickle cell disease, as well as a large proportion of patients with X-linked recessive G6PD deficiency (Fig. 2B). Additionally, a significant proportion of the disorders reported on CTGA are rare, reported in only a handful of families, with ten of these disorders exclusively reported in Emirati patients to date. The presence of multiple cases of rare recessive disorders in consanguineous families affords an opportunity to utilize homozygosity mapping in order to identify disease-causing variants (Maddirevula et al., 2019 ; Alkuraya, 2022). In fact, of the 13 genetic disorders first mapped in Emirati families (Table 1), ten are recessive disorders. As an example, whole genome homozygosity mapping on a family with two cousins affected with a novel recessive phenotype of spondymetaphyseal dysplasia with corneal dystrophy (OMIM# 618961) succeeded in linking the condition to a variant in the *PLCB3* gene (Ben-Salem et al., 2018). A similar approach was used to identify the causal gene in other UAE families, such as *METTL23* in a large inbred UAE family of Yemeni origin with intellectual disability (Reiff et al., 2014) and *SRD5A3* in another UAE family of Baluchi origin with a new type of glycosylation disorder (Cantagrel et al., 2010). However, there are other disorders where the locus has been identified but the gene is still unknown, for example, a new locus for autosomal recessive deafness, DFNB27 (OMIM# 605818) in a large inbred UAE family was identified in 2000 (Pulleyn et al., 2000), but the causative gene has not been identified yet. In the same vein, (al-Gazali et al., 1996) described a branch of a large Bedouin family with multiple children affected with a phenotype similar to McDonough syndrome, in which the gene remains unknown close to three decades later. The presence of familial cases of such disorders within the Emirati population affords a resource that can be used for mapping and localization studies and for understanding the etiology of such conditions.

In order to identify disorders that are rare worldwide, but are relatively over-represented in our study population, we screened our data for rare genetic conditions that have been reported in more than four families (Table 2). Unsurprisingly, the hemoglobinopathies are represented by large groups of patients, reiterating the fact that these disorders cannot be classified as rare in the Emirati population. Type 1 Neurofibromatosis (OMIM# 162200) and Stargardt Disease (OMIM# 248200) are two conditions that have been reported in a sizable number of families. In addition, diseases such as Cystic Fibrosis (OMIM# 219700), Phenylketonuria (OMIM# 261600) and Biotinidase Deficiency (OMIM# 253260) have also been reported in a number of families. These latter diseases are part of the panel of diseases covered by the national newborn screening program, and therefore, affected patients are unlikely to be missed. The national newborn screening program currently covers 19 genetic disorders, including the major hemoglobinopathies, congenital hypothyroidism, congenital adrenal hyperplasia, and some of the amino acid, fatty acid oxidation and organic acid disorders. Interestingly, lysosomal storage disorders, such as Pompe disease (OMIM# 232300) and Alpha Mannosidosis (OMIM# 248500), which are currently absent from the panel, have been reported in more than six Emirati families each in CTGA, indicating that these disorders might warrant an inclusion in then ewborn screening panel.

Founder variants play a major role in defining the genetic landscape of small populations, especially if these populations happen to also be isolated and inbred (Barbouche et al., 2017; Romdhane et al., 2019). We have noted several instances of founder variants propagating through consanguineous families and inbred tribes. In two unrelated Emirati families of Omani origin with *VLDLR*-Associated cerebellar hypoplasia (OMIM# 224050), (Ali et al., 2012) identified the same missense variant, NM_003383.5:c.2117G>T, on a shared haplotype block. Similarly, In five unrelated Emirati fami lies withehlers-Danlos Syndrome, Type VI (OMIM# 225400), the same causal NM_000302.4:c.955C>T variant was seen on a shared haplotype (Giunta et al., 2005). In three separate Emirati tribes, the NM_198535.2:c.436_439del deletion was found to be associated with DECCAGS Syndrome (Saleh et al., 2021). And finally, in a remarkable example, in two UAE tribes, one originating from Oman and one from Yemen, the same NM_001127671.2:c.653dup variant in the *LIFR* gene was found in 50 children from 28 families with Stuve-Wideman syndrome (OMIM# 601559) (Al-Gazali and Hamamy, 2014).

When comparing our Emirati dataset with other databases, we found that 50% and 81% of UAE variants present on CTGA were also present on Clinvar and dbSNP, respectively, while a total of 235 Emirati variants were not available on either (Fig.4). Interestingly, 187 of these variants were reported as being pathogenic or likely pathogenic in the studies where the Emirati subject(s) were described. The lack of ethnic diversity in global genomic databases has repeatedly been raised as a cause of concern (Popejoy and Fullerton, 2016 ; Abou Tayoun and Rehm, 2020; Fatumo et al., 2022). Ethnic and/or national databases like CTGA are crucial in mitigating the under-representation of population-specific variant data. Even within variants reported globally, clinical annotation can vary according to genomic background or with additional data. On CTGA, 55 variants that were reported to be pathogenic or likely pathogenic are currently marked as benign or VUS, or lack a clinical significance on Clinvar. An example of this is the variant NM_003718.5:c.1499C>T in *CDK13*, listed as likely benign on Clinvar but likely pathogenic on CTGA, associated with a congenital syndromic form of cardiac defect (OMIM# 617360). A similar case is the homozygous NM_004453.4:c.807A>C missense variant in the *ETFDH* gene, classified as VUS on Clinvar, which has been reported to be causal for Multiple Acyl-CoA Dehydrogenation Deficiency (OMIM# 231680) in 13 Emirati patients from ten unrelated families. Databases such as CTGA are thus vitally important in providing access to population-based clinical annotation of rare variants. Indeed, analyzing genomes with high runs of homozygosity, known to be rich in Arab-ethnic populations, has provided unique insight into clinically relevant genes (Abouelhoda et al., 2016).

**Figure 4:**
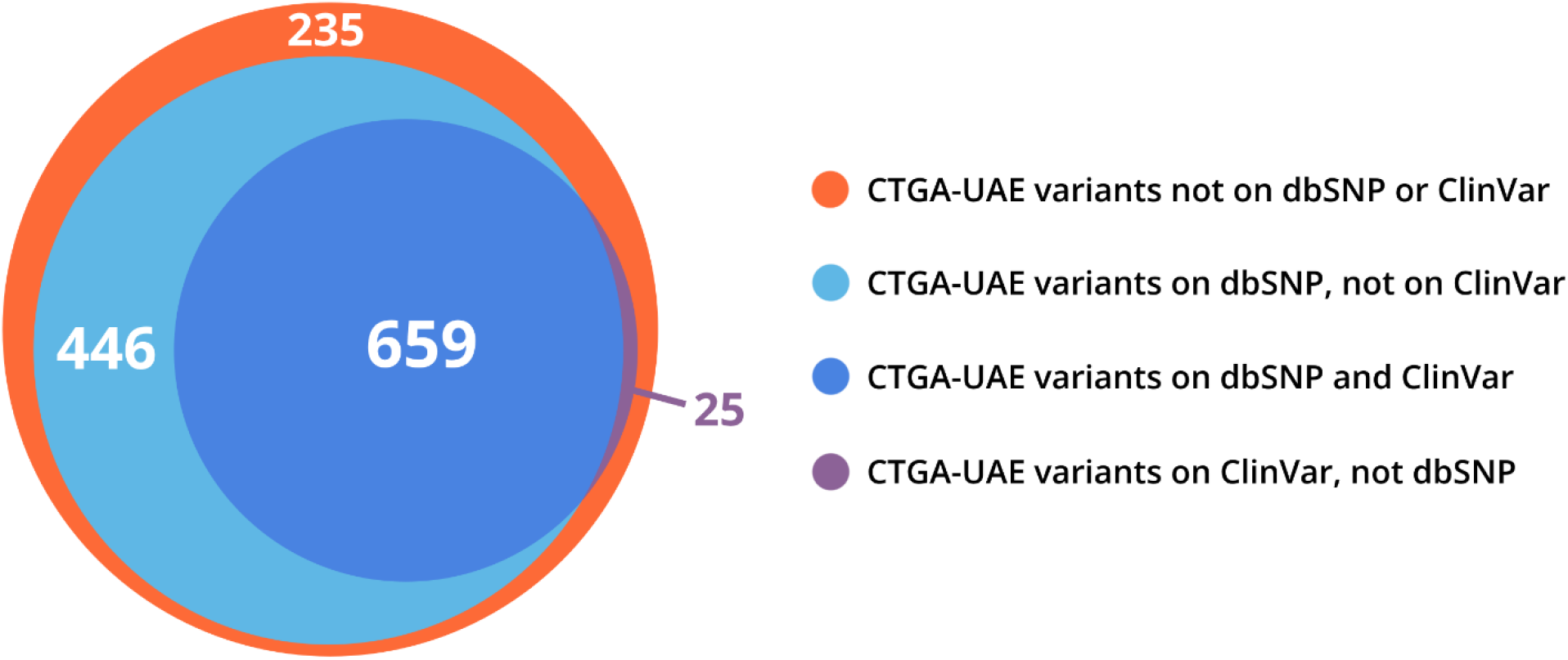
Venn distribution of CTGA variant records reported in Emirati subjects relative to dbSNP and ClinVar variant databases (last updated on 29 December 2022).

The gnomAD and Greater Middle Eastern Variome databases report on aggregate genome and/or exome sequencing data from healthy subjects of various ancestries (Scott et al., 2016). Expectedly, a large number of disease-associated variants on CTGA are not available on either of these datasets. Nevertheless, variants reported as causal in at least one Emirati patient on CTGA were identified on gnomAD (15.4% of variants), gnomAD’s Middle-Eastern subset (1.3%), and GMEV (4.9%). All of these variants exhibited a Minor Alelle Frequency below 5%, further reinforcing the utility of CTGA in describing rare clinically relevant variants.

In conclusion, we provide an overview of the spectrum of genetic conditions that have been described in Emirati individuals along with their associated gene variants. CTGA pools together information from all published reports on genetic disorders and gene variants for Arabs and makes them available for each individual Arab country. This makes it a useful resource for interpreting variants (Sundercombe et al., 2021), especially given ethnic differences, which we have shown examples of in our dataset of Emirati variants. As we have outlined, CTGA contains variant data along with phenotypic data, a feature many other genomic databases lack. This provides clinicians with an opportunity to utilize the database to aid diagnostic and/or therapeutic decisions. Simultaneously, as we have shown, CTGA can also be used to influence screening programs and modify health strategies based on data on both prevalent and rare genetic conditions (Al-Jasmi et al., 2016; Alfadhel et al., 2019). As next generation sequencing technologies get more and more integrated into mainstream healthcare and Emirati genomic data becomes more readily available, we expect the data within CTGA to not only grow, but to also improve publicly available variant annotation, while simultaneously addressing the lack of genomic diversity in global genomic databases.

## Data Availability

All data produced in the present study are available upon reasonable request to the authors. Raw data is also publicly available through the CTGA database.

https://cags.org.ae/en/search-database

## Author Contributions

All authors have made substantial contributions to conception and design of the study and for important intellectual content. SB, PN, SH, AD and SEH have made substantial contributions in acquisition of data, analysis and interpretation of data. SB, PN, SH, AD, MTA, LAG and SEH have been involved in drafting the manuscript and revising it critically. All authors have given final approval of the version to be published.

## Acknowledgments

We thank Andre Megarbane for revising this manuscript.

## Supplementary Material

1. Supplementary Table 1. List of Genetic Phenotypes Reported in Emirati subjects
2. Supplementary Table 2. List of Most Frequently Occurring HPO terms in Emirati Patients with Non-multifactorial Disorders

## Data Availability Statement

The datasets analyzed for this study can be found in the Catalogue for Transmission Genetic in Arabs database, available at https://cags.org.ae/ctga

## References

Abou Tayoun, A.N., Fakhro, K.A., Alsheikh-Ali, A., and Alkuraya, F.S. (2021). Genomic medicine in the Middle East. Genome Med 13(1), 184. doi: 10.1186/s13073-021-01003-9.

Abou Tayoun, A.N., and Rehm, H.L. (2020). Genetic variation in the Middle East-an opportunity to advance the human genetics field. Genome Med 12(1), 116. doi: 10.1186/s13073-020-00821-7.

Abouelhoda, M., Faquih, T., El-Kalioby, M., and Alkuraya, F.S. (2016). Revisiting the morbid genome of Mendelian disorders. Genome Biol 17(1), 235. doi: 10.1186/s13059-016-1102-1.

Al-Ali, M., Osman, W., Tay, G.K., and AlSafar, H.S. (2018). A 1000 Arab genome project to study the Emirati population. J Hum Genet 63(4), 533–536. doi: 10.1038/s10038-017-0402-y.

Al-Gazali, L., and Al i, B.R. (2010). Mutations of a country: a mutation review of single gene disorders in the United Arab Emirates (UAE). Hum Mutat 31(5), 505–520. doi: 10.1002/humu.21232.

Al-Gazali, L., and Hamamy, H. (2014). Consanguinity and Dysmorphology in Arabs. Human Heredity 77(1-4), 93–107. doi: 10.1159/000360421.

al-Gazali, L.I., Aziz, S.A., and Salem, F. (1996). A syndrome of short stature, mental retardation, facial dysmorphism, short webbed neck, skin changes and congenital heart disease. Clin Dysmorphol 5(4), 321–327.

al-Gazali, L.I., Bener, A., Abdulrazzaq, Y.M., Micallef, R., al-Khayat, A.I., and Gaber, T. (1997). Consanguineous marriages in the United Arab Emirates. J Biosoc Sci 29(4), 491–497. doi: 10.1017/s0021932097004914.

Al-Jasmi, F.A., Al-Shamsi, A., Hertecant, J.L., Al-Hamad, S.M., Souid, A.K. (2015). Inborn Errors of Metabolism in the United Arab Emirates: Disorders Detected by Newborn Screening (2011-2014). JIMD Rep. 28:127–135. doi: 10.1007/8904.

Al-Shamsi, A., Hertecant, J.L., Souid, A.K., and Al-Jasmi, F.A. (2016). Whole exome sequencing diagnosis of inborn errors of metabolism and other disorders in United Arab Emirates. Orphanet J Rare Dis 11(1), 94. doi: 10.1186/s13023-016-0474-3.

Alabdullatif, M.A., Al Dhaibani, M.A., Khassawneh, M.Y., and El-Hattab, A.W. (2017). Chromosomal microarray in a highly consanguineous population: diagnostic yield, utility of regions of homozygosity, and novel mutations. Clin Genet 91(4), 616–622. doi: 10.1111/cge.12872.

Alfadhel, M., Umair, M., Almuzzaini, B., Alsaif, S., AlMohaimeed, S.A., Almashary, M.A., Alharbi, W., Alayyar, L., Alasiri, A., Ballow, M., AlAbdulrahman, A., Alaujan, M., Nashabat, M., Al-Odaib, A., Altwaijri, W., Al-Rumayyan, A., Alrifai, M.T., Alfares, A., AlBalwi, M., Tabarki, B. (2019). Targeted SLC19A3 gene sequencing of 3000 Saudi newborn: a pilot study toward newborn screening. Ann Clin Transl Neurol 6(10)2097–2103. doi: 10.1002/acn3.50898.

Ali, B.R., Silhavy, J.L., Gleeson, M.J., Gleeson, J.G., and Al-Gazali, L. (2012). A missense founder mutation in VLDLR is associated with Dysequili brium Syndrome without quadrupedal locomotion. BMC Med Genet 13, 80. doi: 10.1186/1471-2350-13-80.

Aljasmi, F.A., Vijayan, R., Sudalaimuthuasari, N., Souid, A.K., Karuvantevida, N., Almaskari, R., et al. (2020). Genomic Landscape of the Mitochondrial Genome in the United Arab Emirates Native Population. Genes (Basel) 11(8). doi: 10.3390/genes11080876.

Alkuraya, F.S. (2010). Autozygome decoded. Genet Med 12(12), 765–771. doi: 10.1097/GIM.0b013e3181fbfcc4.

Alkuraya, F.S. (2022). Homozygosity mapping: a game-changer for autosomal recessive diseases. Nat Rev Genet 23(1), 2–3. doi: 10.1038/s41576-021-00433-w.

Alsamri, M.T., Alabdouli, A., Alkalbani, A.M., Iram, D., Antony, P., Vijayan, R., et al. (2020). Genetic variants in children with chronic respiratory diseases. Pediatr Pulmonol 55(9), 2389–2401. doi: 10.1002/ppul.24908.

Alshamali, F., Pereira, L., Budowle, B., Poloni, E.S., and Currat, M. (2009). Local population structure in Arabian Peninsula revealed by Y-STR diversity. Hum Hered 68(1), 45–54. doi: 10.1159/000210448.

Arnold Christianson, C.P.H., Bernadette Modell (2006). “March of Dimes Global Report on Birth Defects: The Hidden Toll of Dying and Disabled Children”. (White Plains, New York: March of Dimes Birth Defects Foundation).

Aryasinghe, L., Moezzi, D., Ansari, T., Mathew, E., Sharbatti, S., and Shaikh, R. (2012). Congenital Anomalies at Birth: A Hospital Based Study in UAE. J. Nepal Paediatr. Soc. 32(2), 105–112.

Barbouche, M.R., Mekki, N., Ben-Ali, M., and Ben-Mustapha, I. (2017). Lessons from Genetic Studies of Primary Immunodeficiencies in a Highly Consanguineous Population. Front Immunol 8, 737. doi: 10.3389/fimmu.2017.00737.

Ben-Omran, T., Al Ghanim, K., Yavarna, T., El Akoum, M., Samara, M., Chandra, P., et al. (2020). Effects of consanguinity in a cohort of subjects with certain genetic disorders in Qatar. Mol Genet Genomic Med 8(1), e1051. doi: 10.1002/mgg3.1051.

Ben-Salem, S., Robbins, S.M., Lm Sobreira, N., Lyon, A., Al-Shamsi, A.M., Islam, B.K., et al. (2018). Defect in phosphoinositide signalling through a homozygous variant in PLCB3 causes a new form of spondylometaphyseal dysplasia with corneal dystrophy. J Med Genet 55(2), 122–130. doi: 10.1136/jmedgenet-2017-104827.

Bittles, A.H., and Black, M.L. (2010). Evolution in health and medicine Sackler colloquium: Consanguinity, human evolution, and complex diseases. Proc Natl Acad Sci U S A 107 Suppl 1(Suppl 1), 779–1786. doi: 10.1073/pnas.0906079106.

Bizzari, S., Nair, P., Deepthi, A., Hana, S., Al-Ali, M.T., Megarbane, A., et al. (2021). Catalogue for Transmission Genetics in Arabs (CTGA) Database: Analysing Lebanese Data on Genetic Disorders. Genes (Basel) 12(10). doi: 10.3390/genes12101518.

Bizzari, S., Qari, A., Balobaid, A., Hana, S., Deepthi, A., Nair, P., et al. (2018). “Genetic Disorders in Saudi Arabia: A CTGA Perspective,” in Genetic Disorders in the Arab World: Kingdom of Saudi Arabia, eds. S. EL-HAYEK, M.T.A. ALI & N.A. KHAJA. (Dubai, UAE).

Cantagrel, V., Lefeber, D.J., Ng, B.G., Guan, Z., Silhavy, J.L., Bielas, S.L., et al. (2010). SRD5A3 Is Required for Converting Polyprenol to Dolichol and Is Mutated in a Congenital Glycosylation Disorder. Cell 142(2), 203–217. doi: 10.1016/j.cell.2010.06.001.

CIA (2023). “United Arab Emirates”.

Dawodu, A., Al-Gazali, L., Varady, E., Varghese, M., Nath, K., and Rajan, V. (2005). Genetic contribution to high neonatally lethal malformation rate in the United Arab Emirates. Community Genet 8(1), 31–34. doi: 10.1159/000083335.

Elmusharaf, K., Grafton, D., Jung, J.S., Roberts, E., Al-Farsi, Y., Al Nooh, A.A., et al. (2022). The case for investing in the prevention and control of non-communicable diseases in the six countries of the Gulf Cooperation Council: an economic evaluation. BMJ Glob Health 7(6). doi: 10.1136/bmjgh-2022-008670.

Elsaban, M., Ahmad, D., Toba, N.J., Alsaeed, T., and Abusalah, Z.G. (2021). 400 Five year evaluation of the newborn screening programme in Dubai, United Arab emirates: a cross sectional study. BMJ Paediatrics Open 5(Suppl 1), A116–A117. doi: 10.1136/bmjpo-2021-RCPCH.220.

Fadhil, I., Ali, R., Al-Raisi, S.S., Bin Belaila, B.A., Galadari, S., Javed, A., et al. (2022). Review of National Healthcare Systems in the Gulf Cooperation Council Countries for Noncommunicable Diseases Management. Oman Med J 37(3), e370. doi: 10.5001/omj.2021.96.

Fareed, M., and Afzal, M. (2017). Genetics of consanguinity and inbreeding in health and disease. Ann Hum Biol 44(2), 99–107. doi: 10.1080/03014460.2016.1265148.

Fatumo, S., Chikowore, T., Choudhury, A., Ayub, M., Martin, A.R., and Kuchenbaecker, K. (2022). A roadmap to increase diversity in genomic studies. Nat Med 28(2), 243–250. doi: 10.1038/s41591-021-01672-4.

Giunta, C., Randolph, A., Al-Gazali, L.I., Brunner, H.G., Kraenzlin, M.E., and Steinmann, B. (2005). Nevo syndrome is allelic to the kyphoscoliotic type of the Ehlers-Danlos syndrome (EDS VIA). Am J Med Genet A 133A(2), 158–164. doi: 10.1002/ajmg.a.30529.

Karczewski, K.J., Francioli, L.C., Tiao, G., Cummings, B.B., Alfoldi, J., Wang, Q., et al. (2020). The mutational constraint spectrum quantified from variation in 141,456 humans. Nature 581(7809), 434–443. doi: 10.1038/s41586-020-2308-7.

Koshy, R., Ranawat, A., and Scaria, V. (2017). al mena: a comprehensive resource of human genetic variants integrating genomes and exomes from Arab, Middle Eastern and North African populations. J Hum Genet 62(10), 889–894. doi: 10.1038/jhg.2017.67.

Maddirevula, S., Alzahrani, F., Al-Owain, M., Al Muhaizea, M.A., Kayyali, H.R., AlHashem, A., et al. (2019). Autozygome and high throughput confirmation of disease genes candidacy. Genet Med 21(3), 736–742. doi: 10.1038/s41436-018-0138-x.

Mahfouz, N.A., Kizhakkedath, P., Ibrahim, A., El Naofal, M., Ramaswamy, S., Harilal, D., et al. (2020). Utility of clinical exome sequencing in a complex Emirati pediatric cohort. Comput Struct Biotechnol J 18, 1020–1027. doi: 10.1016/j.csbj.2020.04.013.

Majeed-Saidan, M.A., Ammari, A.N., AlHashem, A.M., Al Rakaf, M.S., Shoukri, M.M., Garne, E., et al. (2015). Effect of consanguinity on birth defects in Saudi women: results from a nested case-control study. Birth Defects Res A Clin Mol Teratol 103(2), 100–104. doi: 10.1002/bdra.23331.

Mills, M.C., and Rahal, C. (2020). The GWAS Diversity Monitor tracks diversity by disease in real time. Nat Genet 52(3), 242–243. doi: 10.1038/s41588-020-0580-y.

MOHAP (2019). “The State of Health Research in the United Arab Emirates”. UAE Ministry of Health and Prevention.

Moundir, A., Ouair, H., Benhsaien, I., Jeddane, L., Rada, N., Amenzoui, N., et al. (2023). Genetic Diagnosis of Inborn Errors of Immunity in an Emerging Country: a Retrospective Study of 216 Moroccan Patients. J Clin Immunol 43(2), 485–494. doi: 10.1007/s10875-022-01398-z.

Patrinos, G.P. (2006). National and ethnic mutation databases: recording populations’ genography. Hum Mutat 27(9), 879–887. doi: 10.1002/humu.20376.

Popejoy, A.B., and Fullerton, S.M. (2016). Genomics is failing o n diversity. Nature 538(7624), 161–164. doi: 10.1038/538161a.

Pulleyn, L.J., Jackson, A.P., Roberts, E., Carridice, A., Muxworthy, C., Houseman, M., et al. (2000). A new locus for autosomal recessive non-syndromal sensorineural hearing impairment (DFNB27) on chromosome 2q23-q31. Eur J Hum Genet 8(12), 991–993. doi: 10.1038/sj.ejhg.5200567.

Reiff, R.E., Ali, B.R., Baron, B., Yu, T.W., Ben-Salem, S., Coulter, M.E., et al. (2014). METTL23, a transcriptional partner of GABPA, is essential for human cognition. Human Molecular Genetics 23(13), 3456–3466. doi: 10.1093/hmg/ddu054.

Romdhane, L., Mezzi, N., Hamdi, Y., El-Kamah, G., Barakat, A., and Abdelhak, S. (2019). Consanguinity and Inbreeding in Health and Disease in North African Populations. Annu Rev Genomics Hum Genet 20, 155–179. doi: 10.1146/annurev-genom-083118-014954.

Saleh, S., Beyyumi, E., Al Kaabi, A., Hertecant, J., Barakat, D., Al Dhaheri, N.S., et al. (2021). Spectrum of neuro-genetic disorders in the United Arab Emirates national population. Clin Genet 100(5), 573–600. doi: 10.1111/cge.14044.

Sanchez-Luquez, K.Y., Carpena, M.X., Karam, S.M., and Tovo-Rodrigues, L. (2022). The contribution of whole-exome sequencing to intellectual disability diagnosis and knowledge of underlying molecular mechanisms: A systematic review and meta-analysis. Mutat Res Rev Mutat Res 790, 108428. doi: 10.1016/j.mrrev.2022.108428.

Scott, E.M., Halees, A., Itan, Y., Spencer, E.G., He, Y., Azab, M.A., et al. (2016). Characterization of Greater Middle Eastern genetic variation for enhanced disease gene discovery. Nat Genet 48(9), 1071–1076. doi: 10.1038/ng.3592.

Sundercombe, S.L., Berbic, M., Evans, C.A., Cliffe, C., Elakis, G., Temple, S.E.L., et al. (2021). Clinically Responsive Genomic Analysis Pipelines: Elements to Improve Detection Rate and Efficiency. J Mol Diagn 23(7), 894–905. doi: 10.1016/j.jmoldx.2021.04.007.

Tay, G.K., Henschel, A., Daw Elbait, G., and Al Safar, H.S. (2020). Genetic Diversity and Low Stratification of the Population of the United Arab Emirates. Front Genet 11, 608. doi: 10.3389/fgene.2020.00608.

Vaisitti, T., Sorbini, M., Callegari, M., Kalantari, S., Bracciama, V., Arruga, F., et al. (2021). Clinical exome sequencing is a powerful tool in the diagnostic flow of monogenic kidney diseases: an Italian experience. J Nephrol 34(5), 1767–1781. doi: 10.1007/s40620-020-00898-8.

Vatsyayan, A., Sharma, P., Gupta, S., Sandhu, S., Venu, S.L., Sharma, V., et al. (2021). DALIA - a comprehensive resource of Disease Alleles in Arab population. PLoS One 16(1), e0244567. doi: 10.1371/journal.pone.0244567.

